# COVID-19 severity is predicted by earlier evidence of accelerated aging

**DOI:** 10.1101/2020.07.10.20147777

**Authors:** Chia-Ling Kuo, Luke C. Pilling, Janice L Atkins, Jane AH Masoli, João Delgado, Christopher Tignanelli, George A Kuchel, David Melzer, Kenneth B Beckman, Morgan E. Levine

## Abstract

With no known treatments or vaccine, COVID-19 presents a major threat, particularly to older adults, who account for the majority of severe illness and deaths. The age-related susceptibility is partly explained by increased comorbidities including dementia and type II diabetes [1]. While it is unclear why these diseases predispose risk, we hypothesize that increased biological age, rather than chronological age, may be driving disease-related trends in COVID-19 severity with age. To test this hypothesis, we applied our previously validated biological age measure (PhenoAge) [2] composed of chronological age and nine clinical chemistry biomarkers to data of 347,751 participants from a large community cohort in the United Kingdom (UK Biobank), recruited between 2006 and 2010. Other data included disease diagnoses (to 2017), mortality data (to 2020), and the UK national COVID-19 test results (to May 31, 2020) [3]. Accelerated aging 10-14 years prior to the start of the COVID-19 pandemic was associated with test positivity (OR=1.15 per 5-year acceleration, 95% CI: 1.08 to 1.21, p=3.2×10^−6^) and all-cause mortality with test-confirmed COVID-19 (OR=1.25, per 5-year acceleration, 95% CI: 1.09 to 1.44, p=0.002) after adjustment for demographics including current chronological age and pre-existing diseases or conditions. The corresponding areas under the curves were 0.669 and 0.803, respectively. Biological aging, as captured by PhenoAge, is a better predictor of COVID-19 severity than chronological age, and may inform risk stratification initiatives, while also elucidating possible underlying mechanisms, particularly those related to inflammaging.

## Introduction

Coronavirus disease 2019 (COVID-19) represents one of the biggest threats to public health in nearly 100 years. While efforts are being undertaken to develop vaccines and antibody tests for COVID-19, in the interim, there is a critical need for assessing risk stratification and to explore the use of geroscience-guided interventions seeking to improve outcomes by targeting biological aging. Accurately identifying those most at-risk of severe complications or death will facilitate treatment decisions and inform guidelines regarding shelter-in-place and social distancing policies. As such, a major priority is in developing biomarkers that prognostically inform on severity of COVID-19 disease progression [5].

The risk of fatality and/or severe complications due to COVID-19 infection is strongly age dependent. On March 18, 2020, the United States Center of Disease Control (CDC) projected that persons ages 85 and older have predicted mortality rates of 10-27%, compared to 3-11% for individuals ages 65-84 years, 1-3% for those 55-64 years, and <1% for those 20-54 years of age [6]. All-in-all, those ages 85 and older have a mortality risk that is 100-fold higher than for those under the age of 50, and currently 8 out of 10 COVID-19 deaths in the United States are among adults age 65 or older. In addition to age, the CDC reports that morbidity prevalence—particularly history of diabetes, cardiovascular disease, chronic kidney disease, liver disease, and chronic obstructive pulmonary disease (COPD)—appears to exacerbate risk of death or symptomatic complications. Similar COVID-19 comorbidities were reported in other countries, e.g., UK [1], China [7], and Italy [8].

Previous studies have predicted COVID-19 outcomes (pneumonia and mortality) using hospital in-patient data including demographics, signs and symptoms, clinical biomarkers, and imaging features. The performance in terms of C-statistic/index or area under the receiver operating characteristic (ROC) curve (AUC) was generally over 90% but subject to bias and overfitting [9]. One study predicted hospital admission related to upper respiratory infections (pneumonia, influenza, acute bronchitis, etc.), proxy events of COVID-19, using over 500 diagnosis features from thousands of general population samples [10]. The resulting AUCs were 70 to 80% but may not be generalized for COVID-19 as risk factors for COVID-19 and for other respiratory infections are not the same [11].

In recent years, we have developed and widely validated several biomarkers of aging [2][12][13] that strongly predict morbidity and mortality risk, in both short-term (1 year) and long-term (25+ years) follow-up [2][13][14]. Based on these observed trends, we hypothesize that biological aging (rather than chronological age) is a strong determinant of symptom severity following COVID-19 infection. We aimed to assess the risk and predictive performance of accelerated aging for COVID-19 severe infection using a biological age measure, named phenotypic age (PhenoAge). PhenoAge was trained using 42 biomarkers as inputs into a supervised machine learning model to predict all-cause mortality [2][14].We applied this measure to biomarker data from 2006 to 2010 of participants from a large community cohort, United Kingdom Biobank (UKB) [15,16]. Combined with information on disease diagnoses updated to 2017, we tested whether PhenoAge was predictive of COVID-19 severity based on mortality data and COVID-19 test results recently linked from the UK National Health Service [3].

## Results

445,875 participants attended baseline assessment centers in England, United Kingdom. Participants who died before the pandemic (set at February 1, 2020, n=24,805) were excluded from our analytical sample. Of the remaining n=421,070 samples (Table 1), 232,184 (55%) were female. 94% of participants self-identified as White (n=393,738), 1.8% identified as Black (n=7,636), and 4.1% identified as Other, which included Mixed, Asian, and Chinese (n=17,307).

**Table 1.** Characteristics of the included samples: participants attending baseline assessment centers in England and alive before the pandemic (set at February 1, 2020)

|  |  |
| --- | --- |
| Included samples | n=421,070 |
| Sex (=female) | 232,184 (55%) |
| Ethnicity |  |
| White | 393,738 (94%) |
| Black | 7,636 (1.82%) |
| Others (incl. Asian, Chinese, and Mixed) | 17,307 (4.13%) |
| Age at baseline (years) | 56.3 ± 8.1 |
| Attained age (to April 26, 2020) (years) | 67.9 ± 8.1 |
| COVID-19 severity groups |  |
| Positive and dead | 197 (0.05%) |
| Positive and alive | 1,273 (0.30%) |
| Negative | 4,630 (1.10%) |
| Untested | 414,970 (98.55%) |
| PhenoAge biomarkers | n=347,751 |
| Albumin (g/L) | 45.28 ± 2.54 |
| Alkaline phosphatase (U/L) | 82.73 ± 22.38 |
| Creatinine (umol/L) | 71.97 ± 14.15 |
| log C-reactive protein (CRP) mg/L | 0.31 ± 1.04 |
| Glucose (mmol/L) | 5.09 ± 0.93 |
| Lymphocyte percentage (%) | 29.03 ± 7.14 |
| Mean corpuscular volume (fL) | 91.03 ± 4.23 |
| Red blood cell distribution width (RDW)(%) | 13.46 ± 0.85 |
| White blood cell count (1000 cells/uL) | 6.83 ± 1.69 |

Among the included samples, 6,100 were tested between March 16 and May 31, 2020. Of the tested samples, 1,273 (20.9%) were positive yet survived, while 197 (3.2%) were positive and died after COVID-19 infection (first death: March 5, 2020, last death: April 26, 2020). The mean attained age on April 26, 2020 (current chronological age) was 67.9 (SD=8.1), where 269,172 (64%) were 65 years and older. PhenoAge at baseline was available for 347,751 of included samples. The mean chronological age (56.3 ± 8.1) was 2.5 years older than the mean PhenoAge (53.8 ± 9.4) at baseline. A summary of the PhenoAge biomarkers at baseline is provided in Table 1.

Two COVID-19 severity outcomes of interest were considered: 1) test positivity (test positive versus the rest, including untested samples and tested negative) and 2) died with test-confirmed COVID-19 (positive dead versus the rest excluding those who were tested positive and survived). Given that viral testing in the study period was largely restricted to symptomatic hospitalized patients (66%), we assume untested samples were enriched for milder or asymptomatic COVID-19 responses. Each severity outcome was used as the dependent variable in the following four logistic models:

M1: current chronological age,

M2: current chronological age + PhenoAgeAccel at baseline,

M3: current chronological age + pre-existing diseases or conditions (“disease states”), and

M4: current chronological age + disease states + PhenoAgeAccel at baseline,

The PhenoAge acceleration (PhenoAgeAccel) was estimated by the residual of PhenoAge adjusted for chronological age at baseline in a linear regression model. As such, PhenoAgeAccel represents how much older (or younger) an individual’s PhenoAge is relative to what is expected based on his/her chronological age. A value of 5 suggests a participant is 5 years older than expected (faster ager), while a value of −5 suggests he/she is 5 years younger than expected (slow ager). Thus, we hypothesize that higher PhenoAgeAccel (faster biological aging) will be positively associated with COVID-19 severity. All the above models were also adjusted for sex, ethnicity, and baseline assessment center in England.

Men were more likely to test positive for COVID-19 and die with test-confirmed COVID-19 than women. Similarly, participants with self-reported black ethnicity were more likely to test positive than those with self-reported white ethnicity, regardless of models (Tables 2 and 3). Current age was minimally associated with test positivity, adjusted for demographics only (M1) or demographics and PhenoAgeAccel assessed 10-14 years prior (M2). However, it was protective when adjusting for pre-existing disease states additionally (M3 and M4). Increased age was associated with increased risk of mortality with or without adjustment for PhenoAgeAccel (M1 and M2), but the effect was moderately reduced with additional adjustment for disease states (M3 and M4).

**Table 2.** Models for COVID-19 test positivity and all-cause mortality with test-confirmed COVID-19: M1 and M2

|  | M1: current chronological age |  | M2: current chronological age + PhenoAgeAccel |  |
| --- | --- | --- | --- | --- |
|  | Positive vs. untested or negative<br>(n=418,681) | Positive dead vs. untested or negative<br>(n=417,418) | Positive vs. untested or negative<br>(n=346,074) | Positive dead vs. untested or negative<br>(n=345,082) |
| Sex<sup>1</sup> (=Male) | **1.33 (1.20, 1.48)**<br>p=5.32e-8 | **2.63 (1.95, 3.57)**<br>p=3.80e-10 | **1.27 (1.13, 1.43)**<br>p=6.63e-5 | **2.18 (1.56, 3.05)**<br>p=5.21e-6 |
| Ethnicity<sup>2</sup> |  |  |  |  |
| Black | **3.47 (2.73, 4.42)**<br>p=4.76e-24 | **5.67 (3.18, 10.10)**<br>p=3.83e-9 | **3.27 (2.48, 4.31)**<br>p=4.80e-17 | **4.92 (2.56, 9.44)**<br>p=1.65e-6 |
| Other (incl. Asian, Chinese, and Mixed) | **2.09 (1.71, 2.57)**<br>p=1.09e-12 | 1.11 (0.51, 2.39)<br>p=0.799 | **2.03 (1.61, 2.56)**<br>p=2.33e-9 | 1.13 (0.49, 2.60)<br>p=0.777 |
| Current Age (per 5 years) | **1.03 (1.00, 1.06)**<br>p=0.075 | **1.86 (1.65, 2.09)**<br>p=3.06e-24 | **1.04 (1.01, 1.08)**<br>p=0.020 | **1.82 (1.60, 2.08)**<br>p=1.19e-19 |
| PhenoAgeAccel (per 5 years) |  |  | **1.30 (1.24, 1.38)**<br>p=1.78e-22 | **1.55 (1.36, 1.76)**<br>p=3.04e-11 |

<sup>1</sup>reference: female; <sup>2</sup>reference: White; in bold if p<0.05; assessment center results skipped

**Table 3.** Models for COVID-19 test positivity and all-cause mortality with test-confirmed COVID-19: M3 and M4

|  | M3: current chronological age + diseases |  | M4: current chronological age + PhenoAgeAccel + diseases |  |
| --- | --- | --- | --- | --- |
|  | Positive alive vs. untested or negative (n=418,644) | Positive dead vs. untested or negative (n=417,381) | Positive alive vs. untested or negative (n=346,055) | Positive dead vs. untested or negative (n=345,063) |
| Sex<sup>1</sup> (=Male) | **1.24 (1.12, 1.38)**<br>p=6.91e-5 | **2.52 (1.84, 3.45)**<br>p=9.95e-9 | 1.23 (1.09, 1.39)<br>p=6.54e-4 | **2.28 (1.61, 3.22)**<br>p=3.42e-6 |
| Ethnicity<sup>2</sup> |  |  |  |  |
| Black | **3.05 (2.39, 3.89)**<br>p=3.62e-19 | **4.37 (2.42, 7.90)**<br>p=1.06e-6 | **3.02 (2.28, 4.00)**<br>p=9.38e-15 | **4.16 (2.13, 8.12)**<br>p=2.87e-5 |
| Other (incl. Asian, Chinese, and Mixed) | **1.90 (1.54, 2.33)**<br>p=1.09e-9 | 0.88 (0.40, 1.92)<br>p=0.747 | **1.89 (1.49, 2.39)**<br>p=1.07e-7 | 0.98 (0.42, 2.29)<br>p=0.970 |
| Current Age (per 5 years) | **0.94 (0.91, 0.98)**<br>p=0.001 | **1.60 (1.41, 1.81)**<br>p=1.25e-13 | **0.97 (0.93, 1.01)**<br>p=0.093 | **1.61 (1.40, 1.84)**<br>p=6.31e-12 |
| PhenoAgeAccel (per 5 years) |  |  | **1.15 (1.08, 1.21)**<br>p=3.20e-6 | **1.25 (1.09, 1.44)**<br>p=0.002 |
| Dementia (46 pos., 13 pos. dead) | **8.56 (6.15, 11.91)**<br>p=3.89e-37 | **8.28 (4.33, 15.83)**<br>p=1.68e-10 | **8.83 (6.14, 12.69)**<br>p=6.59e-32 | **9.14 (4.58, 18.24)**<br>p=3.55e-10 |
| Type II diabetes (201 pos., 51 pos. dead) | **1.61 (1.36, 1.90)**<br>p=2.94e-8 | **2.31 (1.62, 3.30)**<br>p=4.32e-6 | **1.50 (1.24, 1.82)**<br>p=3.51e-5 | **1.94 (1.29, 2.93)**<br>p=0.001 |
| History of pneumonia (124 pos., 23 pos. dead) | **1.95 (1.60, 2.38)**<br>p=3.66e-11 | **1.72 (1.06, 2.78)**<br>p=0.028 | **1.82 (1.45, 2.28)**<br>p=1.73e-7 | 1.44 (0.83, 2.50)<br>p=0.192 |
| Depression (174 pos., 30 pos. dead) | **1.28 (1.09, 1.52)**<br>p=0.003 | **1.90 (1.26, 2.87)**<br>p=0.002 | **1.40 (1.16, 1.67)**<br>p=3.36e-4 | **2.00 (1.28, 3.12)**<br>p=0.002 |
| Atrial fibrillation (126 pos., 28 pos. dead) | **1.61 (1.32, 1.97)**<br>p=2.95e-6 | 1.49 (0.96, 2.29)<br>p=0.073 | **1.56 (1.25, 1.95)**<br>p=8.63e-5 | 1.41 (0.88, 2.28)<br>p=0.153 |
| Hypertension (670 pos., 131 pos. dead) | **1.30 (1.16, 1.46)**<br>p=1.17e-5 | **1.79 (1.29, 2.48)**<br>p=5.09e-4 | **1.25 (1.00, 1.43)**<br>p=9.07e-4 | **1.84 (1.29, 2.63)**<br>p=8.64e-4 |
| COPD (110 pos., 26 pos. dead) | **1.55 (1.25, 1.93)**<br>p=5.54e-5 | **2.04 (1.29, 3.24)**<br>p=0.002 | **1.45 (1.14, 1.85)**<br>p=0.003 | **1.90 (1.14, 3.17)**<br>p=0.013 |
| Chronic kidney disease (55 pos., 9 pos. dead) | **1.93 (1.45, 2.58)**<br>p=7.06e-6 | 1.11 (0.55, 2.27)<br>p=0.769 | **1.77 (1.28, 2.44)**<br>p=5.64e-4 | 0.83 (0.36, 1.89)<br>p=0.657 |
| Liver disease (42 pos., 6 pos. dead) | 1.32 (0.96, 1.82)<br>p=0.083 | 0.96 (0.41, 2.25)<br>p=0.931 | 1.40 (0.99, 1.98)<br>p=0.054 | 0.93 (0.36, 2.39)<br>p=0.878 |
| Rheumatoid Arthritis (38 pos., 12 pos. dead) | 1.11 (0.80, 1.55)<br>p=0.527 | **2.26 (1.23, 4.14)**<br>p=0.008 | 0.99 (0.68, 1.45)<br>p=0.956 | 1.55 (0.74, 3.28)<br>p=0.246 |
| Coronary artery disease (220 pos., 44 pos. dead) | 1.08 (0.92, 1.27)<br>p=0.349 | 0.89 (0.61, 1.29)<br>p=0.539 | 1.09 (0.91, 1.30)<br>p=0.374 | 0.84 (0.56, 1.28)<br>p=0.421 |
| History of delirium (13 pos., 5 pos. dead) | 1.34 (0.73, 2.44)<br>p=0.341 | 1.88 (0.69, 5.14)<br>p=0.216 | 1.30 (0.67, 2.52)<br>p=0.443 | 2.12 (0.75, 6.02)<br>p=0.158 |
| Stroke (46 pos., 10 pos. dead) | 1.19 (0.88, 1.62)<br>p=0.266 | 1.09 (0.56, 2.12)<br>p=0.790 | 1.34 (0.97, 1.85)<br>p=0.077 | 1.29 (0.66, 2.53)<br>p=0.454 |
| Asthma (226 pos., 26 pos. dead) | 0.96 (0.83, 1.12)<br>p=0.607 | 0.71 (0.46, 1.11)<br>p=0.131 | 0.96 (0.81, 1.13)<br>p=0.611 | 0.72 (0.45, 1.16)<br>p=0.175 |
| Previous falls/fragile fractures (387 pos., 65 pos. dead) | 1.08 (0.96, 1.22)<br>p=0.194 | 1.35 (0.99, 1.84)<br>p=0.061 | 1.04 (0.90, 1.19)<br>p=0.613 | 1.20 (0.85, 1.69)<br>p=0.311 |
| Osteoarthritis (175 pos., 35 pos. dead) | 1.02 (0.86, 1.20)<br>p=0.859 | 1.17 (0.80, 1.71)<br>p=0.416 | 0.97 (0.81, 1.17)<br>p=0.759 | 1.16 (0.76, 1.76)<br>p=0.487 |

<sup>1</sup>reference: female; <sup>2</sup>reference: White; in bold if p<0.05; assessment center results skipped

When considering PhenoAgeAccel, the odds ratio per 5-year increase in PhenoAgeAccel was 1.30 (95% CI: 1.24 to 1.38, p=1.78×10^−22^) for test positivity and 1.55 (95% CI: 1.36 to 1.76, p=3.04×10^−11^) for all-cause mortality in M2. This was moderately reduced to 1.15 (95% CI: 1.08 to 1.21, p=3.20×10^−6^) and 1.25 (95% CI: 1.09 to 1.44, p=0.002), with additional adjustment for diseases in M4. The diseases included in M3 and M4 were individually more or less associated with test positivity and all-cause mortality, adjusted for current chronological age, sex, ethnicity, and baseline assessment center in England (Supplementary Table S1). The disease associations tended to be reduced but associations with dementia, type II diabetes, hypertension, and COPD remained statistically significant, with additional adjustment for other diseases only and/or PhenoAge (Table 3). Using samples with complete data to train M1-M4 models, the area under the receiver operating characteristic (ROC) curve (AUC) was 0.669 for test positivity and 0.803 for all-cause mortality using M4, compared to 0.601 and 0.755 using M1 (Figure 1). For the two COVID-19 severity outcomes, the specificity, positive predictive value (PPV), and negative predictive value (NPV) for M1 to M4 when the sensitivity is controlled at 0.6 or 0.8 are presented in Table 4.

**Table 4.**
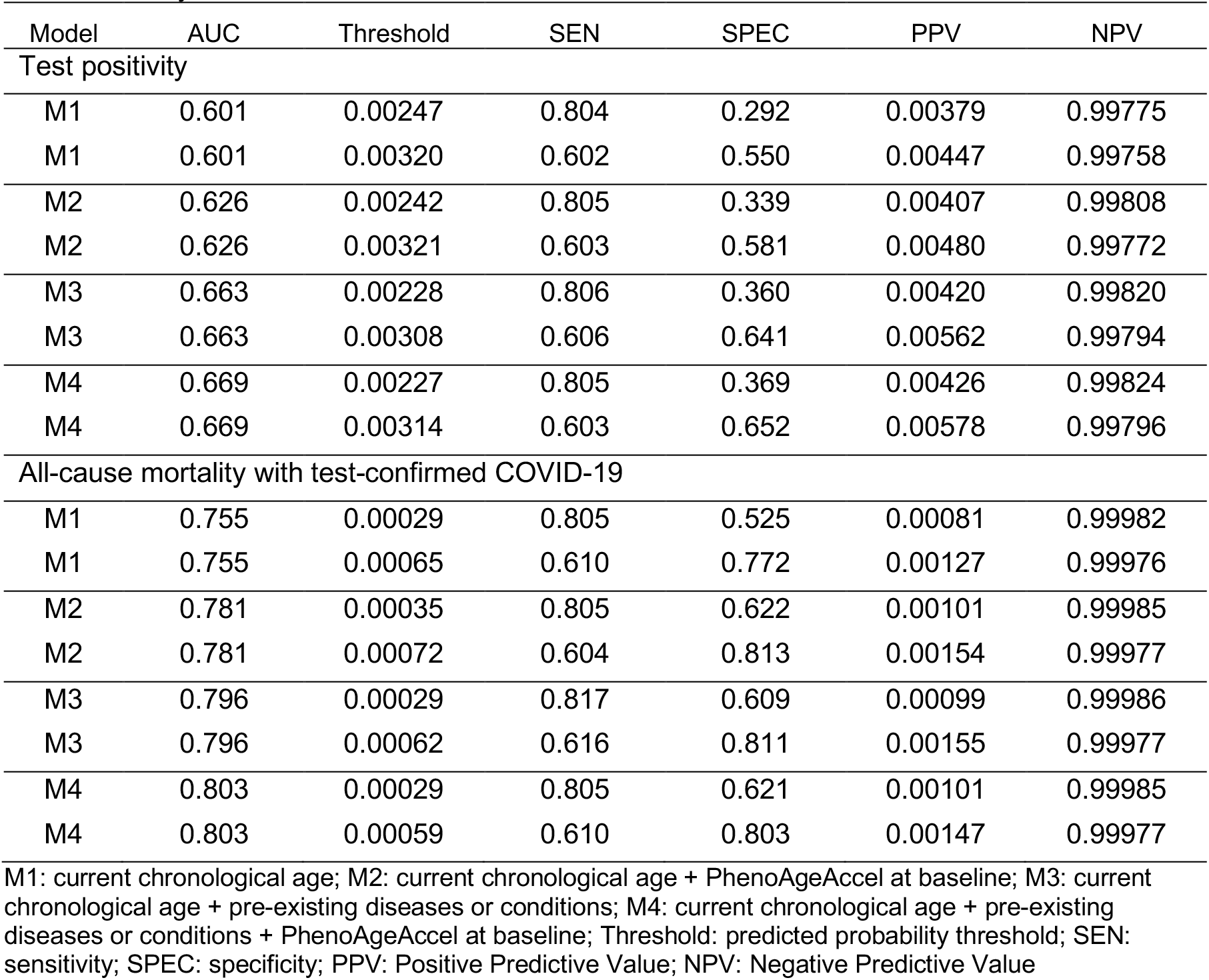
Diagnostic values when the sensitivity is controlled at 0.6, or 0.8 for test positivity or all-cause mortality with test-confirmed COVID-19

| Model | AUC | Threshold | SEN | SPEC | PPV | NPV |
| --- | --- | --- | --- | --- | --- | --- |
| Test positivity |  |  |  |  |  |  |
| M1 | 0.601 | 0.00247 | 0.804 | 0.292 | 0.00379 | 0.99775 |
| M1 | 0.601 | 0.00320 | 0.602 | 0.550 | 0.00447 | 0.99758 |
| M2 | 0.626 | 0.00242 | 0.805 | 0.339 | 0.00407 | 0.99808 |
| M2 | 0.626 | 0.00321 | 0.603 | 0.581 | 0.00480 | 0.99772 |
| M3 | 0.663 | 0.00228 | 0.806 | 0.360 | 0.00420 | 0.99820 |
| M3 | 0.663 | 0.00308 | 0.606 | 0.641 | 0.00562 | 0.99794 |
| M4 | 0.669 | 0.00227 | 0.805 | 0.369 | 0.00426 | 0.99824 |
| M4 | 0.669 | 0.00314 | 0.603 | 0.652 | 0.00578 | 0.99796 |
| All-cause mortality with test-confirmed COVID-19 |  |  |  |  |  |  |
| M1 | 0.755 | 0.00029 | 0.805 | 0.525 | 0.00081 | 0.99982 |
| M1 | 0.755 | 0.00065 | 0.610 | 0.772 | 0.00127 | 0.99976 |
| M2 | 0.781 | 0.00035 | 0.805 | 0.622 | 0.00101 | 0.99985 |
| M2 | 0.781 | 0.00072 | 0.604 | 0.813 | 0.00154 | 0.99977 |
| M3 | 0.796 | 0.00029 | 0.817 | 0.609 | 0.00099 | 0.99986 |
| M3 | 0.796 | 0.00062 | 0.616 | 0.811 | 0.00155 | 0.99977 |
| M4 | 0.803 | 0.00029 | 0.805 | 0.621 | 0.00101 | 0.99985 |
| M4 | 0.803 | 0.00059 | 0.610 | 0.803 | 0.00147 | 0.99977 |

M1: current chronological age; M2: current chronological age + PhenoAgeAccel at baseline; M3: current chronological age + pre-existing diseases or conditions; M4: current chronological age + pre-existing diseases or conditions + PhenoAgeAccel at baseline; Threshold: predicted probability threshold; SEN: sensitivity; SPEC: specificity; PPV: Positive Predictive Value; NPV: Negative Predictive Value

**Figure 1.**
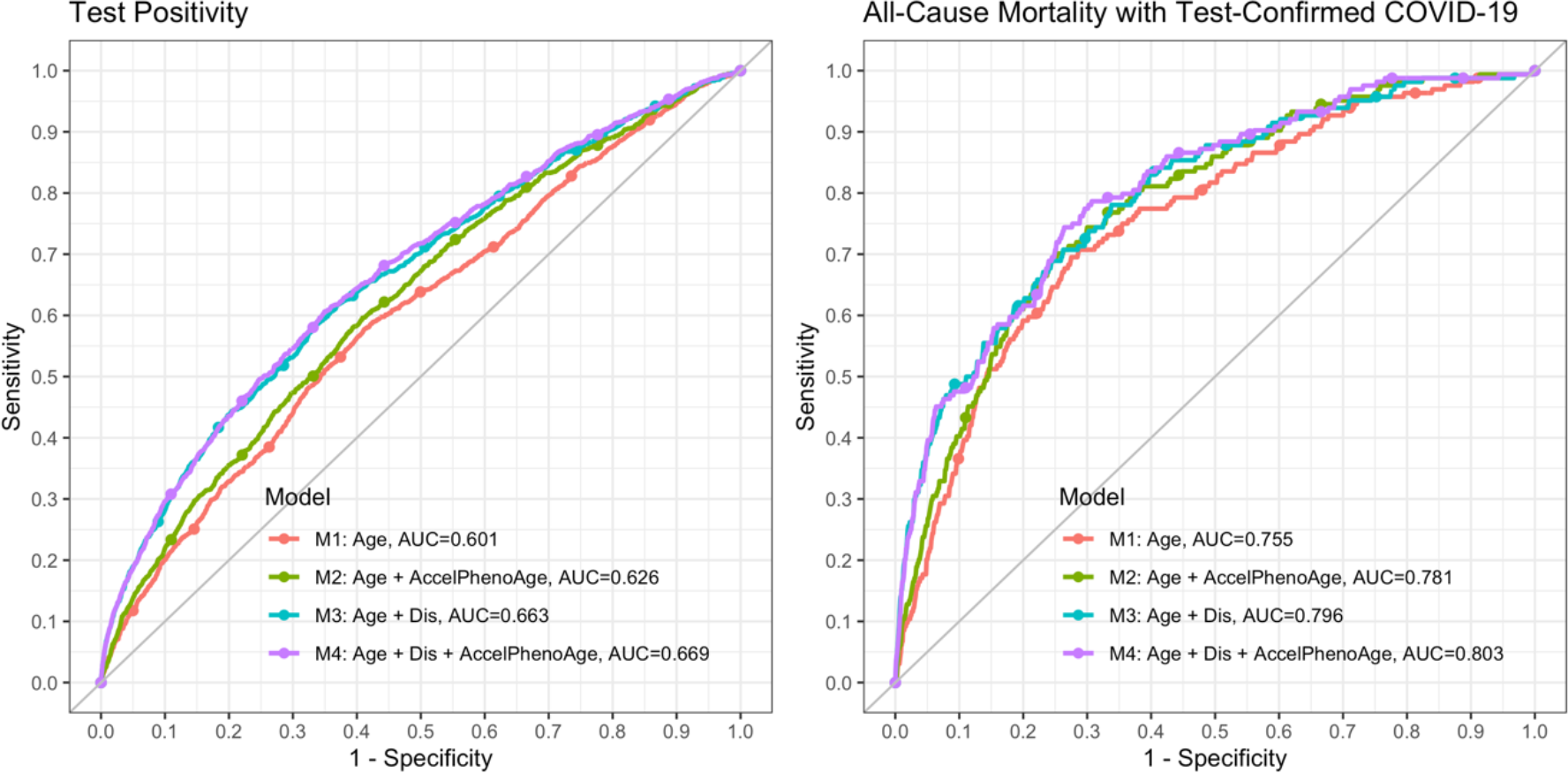
Receiver operating characteristic curves (ROCs) and areas under the ROC curves (AUCs) for test positivity and all-cause mortality with test-confirmed COVID-19

The sensitivity and PPV are probably the most important diagnostics here, representing percent of severe cases that will be pre-identified (sensitivity) and percent of identified cases that will be severely infected (PPV). A subject is identified for high risk of severe infection if the predicted probability is greater than the threshold for a desired sensitivity. For test positivity, 426 per 100,000 identified cases are expected to be tested positive or hospitalized (PPV=426/100,000 versus the sample test positivity rate 349 per 100,000) using M4 with the predicted probability threshold 0.00227 for 80% sensitivity, and the PPV increases to 578/100,000 when the sensitivity is set to 0.6. For all-cause mortality, 101 per 100,000 identified cases are expected to die with test-confirmed COVID-19 (PPV=101/100,000 versus the sample all-cause mortality rate 47 per 100,000) using M4 with the predicted probability threshold 0.00029 for 80% sensitivity, and the PPV increases to 147/100,000 when the sensitivity is set to 0.6.

For sensitivity analysis, we modelled the nine PhenoAge biomarkers jointly instead of PhenoAgeAccel in M2. Multiple biomarkers were associated with test positivity (p<0.05, Figure 2): the strongest association appeared to be with albumin (OR=0.87 per SD increase in albumin, 95% CI: 0.82 to 0.93, p=1.01×10^−5^), yet other significant biomarkers included glucose (OR=1.09, 95% CI: 1.04 to1.14, p=0.001), log(CRP) (OR=1.10, 95% CI: 1.04 to 1.18, p=0.002), RDW (OR=1.09, 95% CI: 1.03 to 1.16, p=0.002), white blood cell count (OR=1.09, 95% CI: 1.03 to 1.16, p=0.003), and creatinine (OR=1.10, 95% CI: 1.03 to 1.18, p=0.004). The associations between the biomarkers and test positivity or all-cause mortality were consistent (Figure 2). The associations with all-cause mortality tended to be underpowered as the COVID-19 mortality rate is extremely low. Still, glucose (OR=1.21 per SD increase in glucose, 95% CI: 1.09 to 1.34, p=2.31×10^−4^) was significantly associated with all-cause mortality, as well as log (CRP)(OR=1.20, 95% CI: 1.01 to 1.42, p=0.037) and RDW (OR=1.18, 95% CI: 1.02 to 1.37, p=0.027). The AUC for M2 with either PhenoAgeAccel or the nine biomarkers was similar for test positivity (0.626 versus 0.632) and for all-cause mortality with test-confirmed COVID-19 (0.781 versus 0.777).

**Figure 2.**
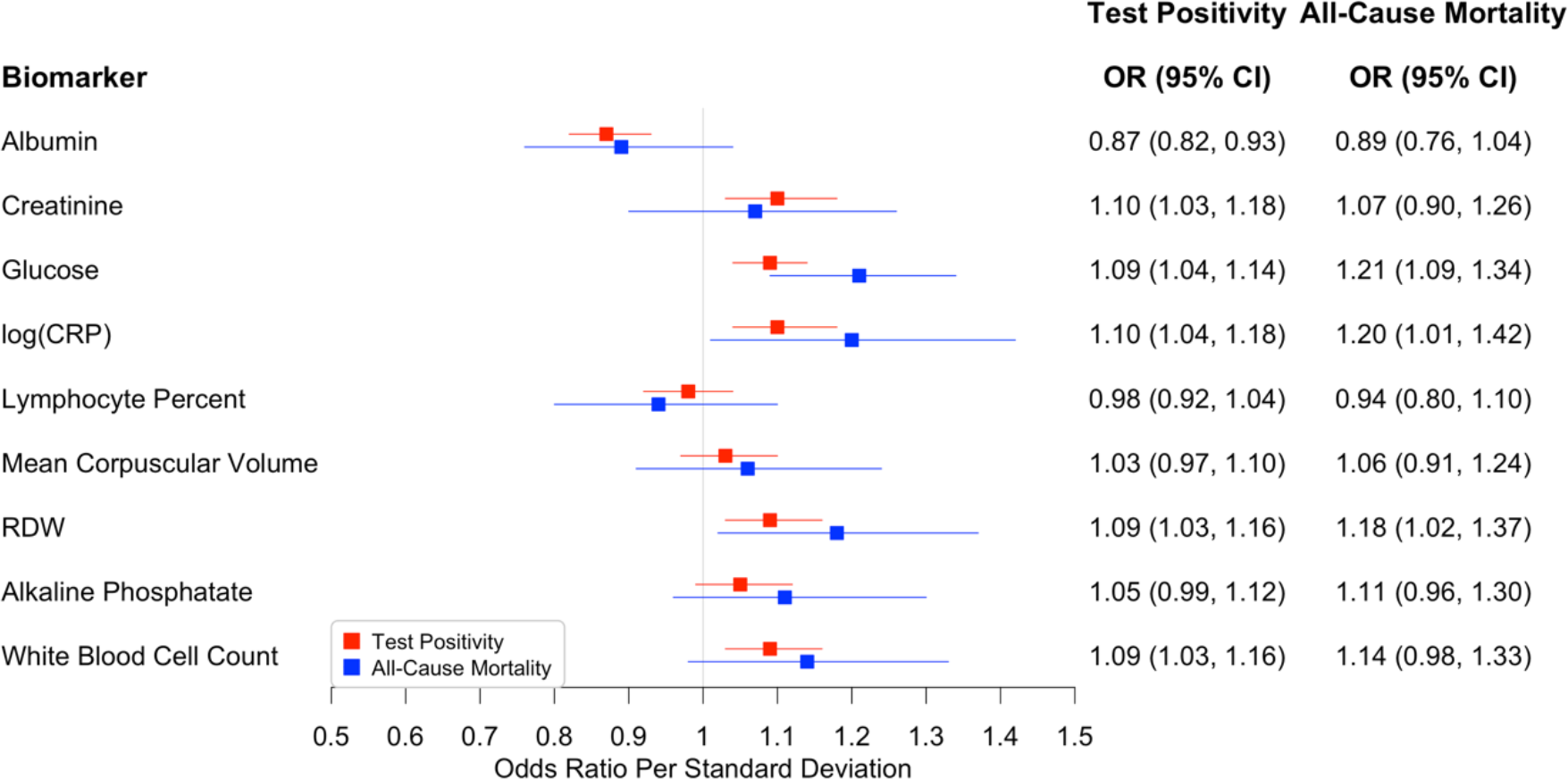
Associations between the nine PhenoAge biomarkers and test positivity or all-cause mortality with test-confirmed COVID-19, with adjustment for age at baseline, sex, ethnicity, and baseline assessment center in England

We also built M1-M4 for inpatient test positivity comparing inpatient positives (n=975, 66% of 1470 positives in total) to untested samples and those tested negative. The results were similar to those for test positivity (Supplementary Tables S2 and S3), e.g., the odds ratio for inpatient test positivity was 1.15 (95% CI: 1.07 to 1.23) per 5-year increase in accelerated PhenoAge versus 1.15 (95% CI: 1.08 to 1.21) for test positivity, using the model M4. Current chronological age was more strongly associated with inpatient test positivity than with test positivity, when diseases were not adjusted. The AUC for inpatient test positivity was 0.682 versus 0.669 for test positivity using M4. As for test positivity, albumin (OR=0.86, 95% CI: 0.80 to 0.92 per SD increase in albumin) was more strongly associated with inpatient test positivity than other PhenoAge biomarkers adjusting for sex, ethnicity, current chronological age, and baseline assessment center.

## Discussion

Accelerated aging measured by PhenoAge more than a decade prior to the pandemic was associated with increased COVID-19 symptom severity, as measured by test positivity and all-cause mortality with test-confirmed COVID-19. These associations were significant even when adjusting for various chronic diseases simultaneously. Similarly, some disease associations were lessened with the addition of PhenoAgeAccel (M3 vs. M4), including associations with type II diabetes, COPD, history of pneumonia, and chronic kidney disease. The association with COVID-19 severity that these diseases share with PhenoAge may reflect the role of underlying immune-related pathways. In our recent genome-wide association study (GWAS) on accelerated PhenoAgeAccel, we observed enrichment for biological processes involved in immune system, cell function, and carbohydrate homeostasis [17]. A methylation clock (DNAmPhenoAge) trained using PhenoAge as a surrogate for biological age, instead of chronological age, has been shown to be associated with activation of pro-inflammatory, interferon, DNAm damage repair, transcriptional/translational signaling, and various markers of immunosenescence: a decline of naïve T cells and shortened leukocyte telomere length [2]. Overall, this suggests that the accelerate biological aging profile captured by PhenoAge is largely characterized by accelerated inflammaging (chronic low-grade inflammation) [18], whose underlying mechanisms may contribute to severe COVID-19 symptoms. These fundamental biological aging processes—including, genomic instability, cell senescence, mitochondria dysfunction, microbiota composition changes, NLRP3 inflammasome activation, primary dysregulation of immune cells, and chronic infections—may serve as potential targets for prevention of severe infection of COVID-19 [19].

The effect of current chronological age on test positivity or all-cause mortality following COVID-19 infection only slightly changed with additional adjustment for PhenoAgeAccel. This likely reflects the variance in timing between the initial blood draw used to estimate PhenoAge and the COVID-19 pandemic. The length of time that participants were followed remains an independent predictor, with longer follow-up associated with older ages and increased COVID-19 severity. We hypothesize that had PhenoAgeAccel been assessed simultaneously for all participants, the effect of chronological age may have been diminished or even reversed in a manner consistent with our previous findings for all-cause and disease-specific mortality [14]. In fact, reversal of the chronological age association—increased age tended to reduce the likelihood of test positivity— was seen when adjusting for disease states. One explanation is that among people with comparable disease states, having a younger chronological age may reflect faster biological aging in those individuals. Nevertheless, the effect of age on mortality remained strong after adjustment, which could suggest that disease state alone is not sufficient to capture the processes underlying the strong force of mortality in advanced age in this young cohort.

Most notable, perhaps, is our finding that an integrative measure assessed in individuals a decade or longer prior to the current outbreak – divorced from chronological age and disease states – can provide such strong predictive power. The biomarkers in PhenoAge were selected based on their joint ability to predict age-related mortality. The biomarker-specific associations in this study, may shed light on the key aging signals that may be driving the association between biological aging and COVID-19 severity. We found that multiple biomarkers in PhenoAge were associated with COVID-19 severity, with the strongest associations found between albumin and test positivity and between glucose and all-cause mortality with test-confirmed COVID-19. Previous studies [20][21][22] have linked albumin to COVID-19 severity in hospitalized patients. We replicated the association using albumin levels assessed more than a decade before the pandemic, supporting the hypothesis that hypoalbuminemia is not due to decreased albumin synthesis in severely infected patients but chronic inflammation [22], also indicated by elevated CRP and white blood cell count. The association between glucose and all-cause mortality was not surprising as diabetes is a leading cause of COVID-19 mortality [23]. Elevated levels of glucose may increase viral replication in vivo and suppress the anti-viral immune response. Hyperglycemia has been linked to fatal outcomes in patients infected with influenza [24], which is consistent with our finding on glucose and all-cause mortality following COVID-19 infection.

PhenoAges were estimated from blood draws that also took place more than a decade before the pandemic arose. On one hand, it is suggesting that an aging measure dating back that far had bearing on contemporaneous disease risk. This also provides further evidence that the PhenoAge and COVID-19 severity association is not completely due to underlying pathology or presence of disease. However, we hypothesize that some people who appeared to be “fast agers” at the time of blood draw may have improved their health, while others may have experienced worsening health over the decade. If so, we would expect the prognostic value of this measure to be enhanced by a more recent estimate of PhenoAge.

One way to treat COVID-19 is to target the underlying mechanism. In this perspective, we have suggested that inflammation-related pathways may be potential targets due to the biological implication from the association between accelerated PhenoAge and COVID-19 severity. Alternatively, COVID-19 may be treated by reversing the aging process as the case fatality rate, increases with chronological age, PhenoAge, and burden of age-related diseases. Drugs under development to slow the aging process, e.g., rapamycin and metformin, may be used to treat COVID-19, building of the hypothesis and our results showing that people with younger biological ages are less prone to age-related diseases [4]. Rapamycin has been shown to slow aging in a number of model organisms and in humans and it was shown to increase the effectiveness of the influenza vaccine [25]. Additionally, metformin is known for lowering blood glucose levels, which are associated with COVID-19 severity, and there is new evidence mounting to suggest that metformin also has the potential to inhibit the pro-inflammatory phenotype of immune cells [26]. Overall, our findings further substantiate the call for trials on longevity drugs to treat and/or prevent COVID-19 severity and lethality.

There are limitations to this study, which warrant acknowledgement. First, the disease status was determined based on self-reported doctor diagnoses at baseline and hospital admission records to 2017 (3 years before the pandemic), without use of the primary care data (to 2017, for nearly half of the UKB participants). Second, we did not include cancers in the analysis as the status of cancer (e.g., progressing vs. remission) is not available, which is strongly associated with mortality related to COVID-19 [27]. Additionally, some participants are not old enough to develop late-onset diseases. As disease cases may be misclassified as non-disease cases, the disease odds ratio estimates were likely to be biased towards the null [28]. Also, clinical severity data is not available, but we used the mortality data to derive the severity outcome, all-cause mortality with test-confirmed COVID-19. While the mortality data is incomplete (censored at March 31, 2020, with additional mortality data from April, 2020), we excluded those who were tested positive and survived so the impact on the results of all-cause mortality should be minimal. Lastly, the UKB sample is known to be healthier than the general population [29]; however, risk factor associations are usually generalizable [30].

In conclusion, accelerated aging measured by PhenoAge was associated with both COVID-19 severity outcomes, with adjustment for demographics including current chronological age, and disease comorbidities. Accelerated PhenoAge in combination with demographics and disease comorbidities produced positive predicted values greater than the sample test positivity rate and all-cause mortality rate after COVID-19 infection. Accelerated aging by PhenoAge is largely characterized by inflammaging, suggested by the composition of biomarkers, the shared association with COVID-19 severity between PhenoAge and multiple diseases, and our prior gene enrichment analysis results of accelerated PhenoAge. Targeting the potential mechanisms underlying inflammaging may reduce COVID-19 severity.

## Materials and Methods

### UK Biobank data

United Kingdom Biobank (UKB) [15,16] recruited over 500,000 subjects between the ages of 40 and 70 during 2006 to 2010. We restricted analyses to participants attending baseline assessment centers in England, excluding those who died before February, 2020. At recruitment (baseline), biological samples of participants were collected for biomarker assays. The disease status was confirmed based on self-reported doctor diagnoses at baseline or hospital admission records updated to March, 2017. Also, the mortality data was used based on death certificates, censored at March 31, 2020, plus incomplete mortality data from April, 2020. These phenotypic data are linked to the UK national COVID-19 test results, currently from March 16 to May 31, covering the peak of COVID-19 incidence.

Two COVID-19 severity outcomes were created: test positivity (versus the rest including untested samples and tested negative) and all-cause mortality with test-confirmed COVID-19 (versus the rest excluding those who were tested positive and survived). Test positivity is a proxy for COVID-19 severity as testing during the above period was largely restricted to hospital in-patients with clinical signs of infection [3].

### PhenoAge

PhenoAge [2] is a biological age measure in the same scale as that of chronological age. An excessive PhenoAge compared to the chronological age suggests accelerated aging. PhenoAge was developed based on mortality scores from the Gompertz proportional hazard model on chronological age and nine biomarkers (Table 1), which were selected from 42 biomarkers by Cox penalized regression model for best predicting mortality in the National Health and Nutrition Examination Survey (NHANES) III [2].

Biomarkers in UKB were measured at baseline for all participants (measurement details in the UK Biobank Biomarker Panel [31] and UK Biobank Haematology Data Companion Document [32]). To correct distribution skewness, we set the top and bottom 1% of values to the 99^th^ and 1^st^ percentiles. The formula of PhenoAge is given by

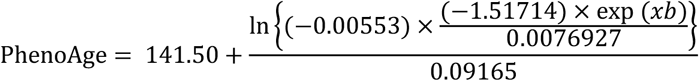

where

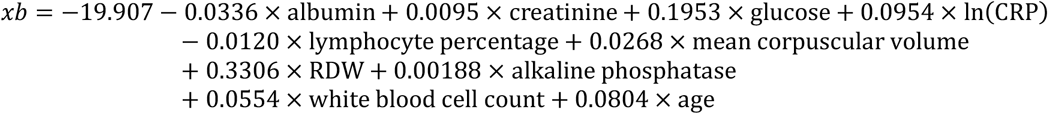

and denotes the chronological age.

### Statistical Methods

We linked the two COVID-19 severity outcomes to PhenoAge acceleration (PhenoAgeAccel), demographics including current chronological age (set at April 26, 2020, when the last death occurred in the data), and disease comorbidities by four logistic models:

M1: current chronological age

M2: current chronological age + PhenoAgeAccel at baseline

M3: current chronological age + pre-existing diseases or conditions

M4: current chronological age + pre-existing diseases or conditions + PhenoAgeAccel at baseline

where PhenoAgeAccel was estimated by the residual from the linear regression model of PhenoAge on chronological age at baseline. The above models were adjusted for sex, ethnicity, and baseline UKB assessment center in England to account for geographic differences in the prevalence of COVID-19. The pre-existing diseases or conditions were selected as those included in Atkins et al. [1] and liver disease (ICD-10 codes in Supplementary Table 4): dementia, type II diabetes, history of pneumonia, depression, atrial fibrillation, hypertension, COPD, chronic kidney disease, rheumatoid arthritis, coronary artery disease, history of delirium, stroke, asthma previous falls/fragile fractures, and osteoarthritis.

We also evaluated M1-M4 for discriminative power using 10-fold cross validation by the area under the ROC curve (AUC). Specificity, positive and negative predictive values were reported when the sensitivity was controlled at 0.6 or 0.8 by manipulating the predicted probability threshold to predict who will be severely infected.

For sensitivity analysis, we restricted test positivity to inpatient test positivity by origins when the sample was taken and excluded non-inpatient positives. We also replaced PhenoAgeAccel in M2 by the nine individual PhenoAge biomarkers at baseline (M2Biomarkers),

M2Biomarkers: current chronological age + glucose + ln(CRP) + lymphocyte percentage + mean corpuscular volume + RDW + alkaline phosphatase + white blood cell count where each biomarker was z-transformed to be in the same scale for the effect comparison with other biomarkers, within and between models. Additionally, sex, self-reported ethnicity, and assessment center at baseline were adjusted. All the statistical analyses were performed in R version 3.4.1.

## Data Availability

This research was conducted using the UK Biobank resource, under the application 14631.

## Acknowledgments

The authors are supported by grants funded by the National Institute on Aging, National Institute of Health: R00AG052604 for CLK and MEL; R21AG060018 for CLK, LCP, GAK, and DM; R33AG061456 for GAK. DM and LCP are supported by the University of Exeter Medical School, and in part by the University of Connecticut School of Medicine. JLA is funded by the UK Medical Research Council (MR/S009892/1). JM is funded by the National Institute for Health Research (NIHR), (NIHR Doctoral Research Fellowship, DRF-2014-07-177). UK Biobank received an approval from the UK Biobank Research Ethics Committee (REC; REC reference 11/NW/0382). All the participants provided written informed consent to participate in the study and for their data to be used in future research. This research was conducted using the UK Biobank resource, under the application 14631. The views expressed in this publication are those of the author(s) and not necessarily those of the NHS, the National Institute for Health Research or the Department of Health and social care.

## Notes

### Competing Interest Statement

The authors have declared no competing interest.

### Author Declarations

Project is not human subject research and IRB involvement is not required.

